# Environmental Influences on Childhood Asthma Prevalence in Philadelphia

**DOI:** 10.1101/2021.06.08.21258548

**Authors:** Raeva N. Mulloth, Alexander J Blackley, Peter J. Koszuta, Kaitlyn M. Nemes, Maddison M. Vail, Monglin L. Zhang

## Abstract

**Background:** In 2019, the American Lung Association found that, for the second year in a row, the Philadelphia metro has worsened the surrounding area’s air quality, due to worsening ozone smog. This spike in unhealthy air quality in Philadelphia has affected the health of the city’s population. Unhealthy air quality can be exacerbated by asbestos, which has been found in many Philadelphia elementary schools. Although asbestos usage is now highly regulated, it can still be found in consumer products and construction material today. Among the many factors contributing to asthma onset and other lung diseases, air pollution and dangerous air particles such as asbestos are important contributors. Children in these asbestos infected schools became exposed and ultimately sick which led to their school’s eventual closure. Due to children’s immature and more vulnerable airways, this exposure may have led to increasing cases of respiratory distress.

**Methods:** This research study analyzed publicly available asbestos data from Asbestos Hazard Emergency Response Act (AHERA) reports from four Philadelphia elementary schools (Laura H. Carnell, James J. Sullivan, Clara Barton, and Thomas M. Peirce) from 2016-2018 to further understand the influence of asbestos particles on asthma in children. Secondary data analysis determined the levels of asbestos contamination in each elementary school and the severity of the condition for each school. This was compared to children’s asthma prevalence during the selected time period.

**Results:** Asbestos was mainly found in the 2-6 inch pipe insulation and tiles within each school. Between 0.06 and 1.18% asbestos damage was found in 2-6inch pipe insulation in schools closed for asbestos abatement. An r^2^ of 0.9997 was found when comparing the 206inch pipe damage percentage and the newly friable material found in each school. Thomas M. Pierce Elementary was determined to be the highest concern according to the analysis of the AHERA reports.

**Conclusion:** Children exposed to asbestos in elementary schools, and with a predisposition to asthma, were more likely to suffer from respiratory distress, due to the multiple contributing environmental factors.

## Introduction

The City of Philadelphia Department of Public Health Air Management Services has been tracking air quality since the 1980s and discovered an increase in ozone smog in recent years that has contributed to unhealthy air quality (1). Oxidative species exposure, such as ozone and particulate matter found in the air, can lead to tougher, more fibrous, and less efficient respiratory tissue. These environmental influences put those with existing respiratory conditions and vulnerable populations, such as elderly and developing children, at a higher risk for respiratory distress (1,3). Asthmatic children who have been exposed to ozone-polluted air can be more at risk for hospitalization, and as a consequence, live shorter lives (2,4). Negative outcomes related to respiratory illness in Philadelphia can be a results of ozone levels as well as other air pollutants (4).

Children are especially vulnerable to respiratory irritation and distress due to their behavior, and physiology, specifically the immaturity of their airways (2). Evidence supports that air pollution in developing cells early in life leads to abnormal respiratory functions and growth, which leads to the development of respiratory diseases later on in life (2,5). Another environmental factor that influences Philadelphia children’s propensity for respiratory diseases is their exposure to asbestos-infected schools. Asbestos is a naturally occurring mineral that is resistant to heat, electricity, and corrosion, making it an effective insulator (6). From 1866-1978, asbestos-containing building material was manufactured because it was considered to be the ideal material for all types of insulations. Although asbestos presented as a great insulator it is highly toxic with prolonged exposure (6). Buildings built before asbestos was deemed toxic were all insulated with asbestos including many of the elementary schools in Philadelphia (7). Data was collected from different elementary schools in Philadelphia from 2016-2018 that were built between 1908 and 1931. These schools have confirmed findings of asbestos in their pipe and duct installation, and floor tiles (7–10). The asbestos in these elementary schools potentially has exposed a total of 2,700 students which may be associated with an increase in incidence rates of asthma in these children due to the toxicity of the product (7–10).

In addition to assessing asbestos within the identified elementary schools, other information related to air quality and respiratory illness in Philadelphia was collected. Data collected from the city of Philadelphia showed rates of asthma in specific neighborhoods in regard to population density and location. Research from 2019, conveys the breakdown of 46 different neighborhoods within Philadelphia and compares averages of many different health factors and health outcomes within each neighborhood and in Philadelphia (11). The data shows that neighborhoods in Philadelphia with higher asthma rates in children are located in the Upper North, Lower Northeast, and West areas (11). The neighborhoods and surrounding areas of this study’s chosen elementary schools have an increased prevalence of childhood asthma compared to other Philadelphia neighborhoods (11). These neighborhoods also have reported differing levels of access to medical care and housing code violations, which are factors that could contribute to poorer health outcomes for children with asthma. Therefore, it is imperative to understand the harm that asbestos can cause in these areas. Our study characterized children from asbestos infected schools in Philadelphia with their negative health outcomes (asthma prevalence).

The goal of this research is to bring awareness to this rising concern, as to eventually finance the reconstruction of the many asbestos infected schools in Philadelphia in order to provide better air quality for young children with asthma.

## Methods

### Participants

This study utilized de-identified patient data from the Philadelphia Department of Public Health. This information was supplemental data about the overall health and asthma prevalence in Philadelphia neighborhoods where these schools are located.

This report mainly used the Philadelphia Asbestos Hazard Emergency Response act (AHERA) collective reports on inspection details for asbestos management plans from different elementary schools. The schools looked at were Laura H. Carnell Elementary School (“Carnell”), James J. Sullivan Elementary School (“Sullivan”), Thomas M. Peirce Elementary School (“Pierce”), and Clara Barton Elementary School (“Barton”). The time frame was between 2016 and early 2019, due to available data in AHERA reports (7–10). These schools served as a foundation to the study of asthma in children and could support the idea of asbestos exposure increasing incidence of asthma. Information was gathered about confirmed asbestos findings in the school’s building materials through room-by-room inspection by a professional general contractor (9). From these inspection plans, this study extracted what building materials were of high asbestos content noting location, type, amount present, amount damaged, damage score, and plan of action. Based on this data and the school’s average class sizes, the total number of students that had been exposed to asbestos was determined. The student population was estimated to be around 2,700 students, which was reported in the school’s demographic sheets.

This study was determined by the Geisinger IRB as research that does not involve or interact with human subjects.

### Data Analysis

Secondary data and reports were analyzed to acknowledge the instance of poor air quality, high asbestos levels, and higher rates of asthma in children. Our data collection methods started by looking at AHERA data reports about the inspections documenting asbestos levels (7–9). Each report was analyzed by extracting data by noting the amount of material, amount of damage, damage potential, and response actions, in each elementary school room. We analyzed where asbestos was found inside of the schools and determined if asbestos was in high traffic. Classrooms, hallways, nurses office, large classroom closets, cafeteria and kitchen, were all areas included in data analysis since children could be expected to interact in these environments. The asbestos could be found in either floor tiling, pipe insulation, or duct insulation. Utilizing PRISM, data on how much asbestos was found in these areas, where it was present, and the condition, was graphed.

Incident rate reports from Philadelphia Department of Public Health for asthma in children, as well as the health outcomes and comparative asthma rates of the surrounding neighborhoods were analyzed. Environmental measures, such as volatile organic compounds (VOC) and ozone levels, were collected from data published by the Department of Public Health in the city of Philadelphia (1). Data points were analyzed in geographic groups around elementary schools and neighborhoods of metropolitan Philadelphia. Comparisons made between these geographic groups and demographics included the age of patients with signs of asthma, elementary school exposure to asbestos, and the effect of environmental air quality.

## Results

Figure 1 shows the prevalence of childhood asthma in the United States (US) and Pennsylvania (PA). Since 2000, PA has had a higher number of children with asthma compared to the rest of the nation. As of 2016, the rate is steadily increasing in PA, while in the US it is decreasing.

**Figure 1:**
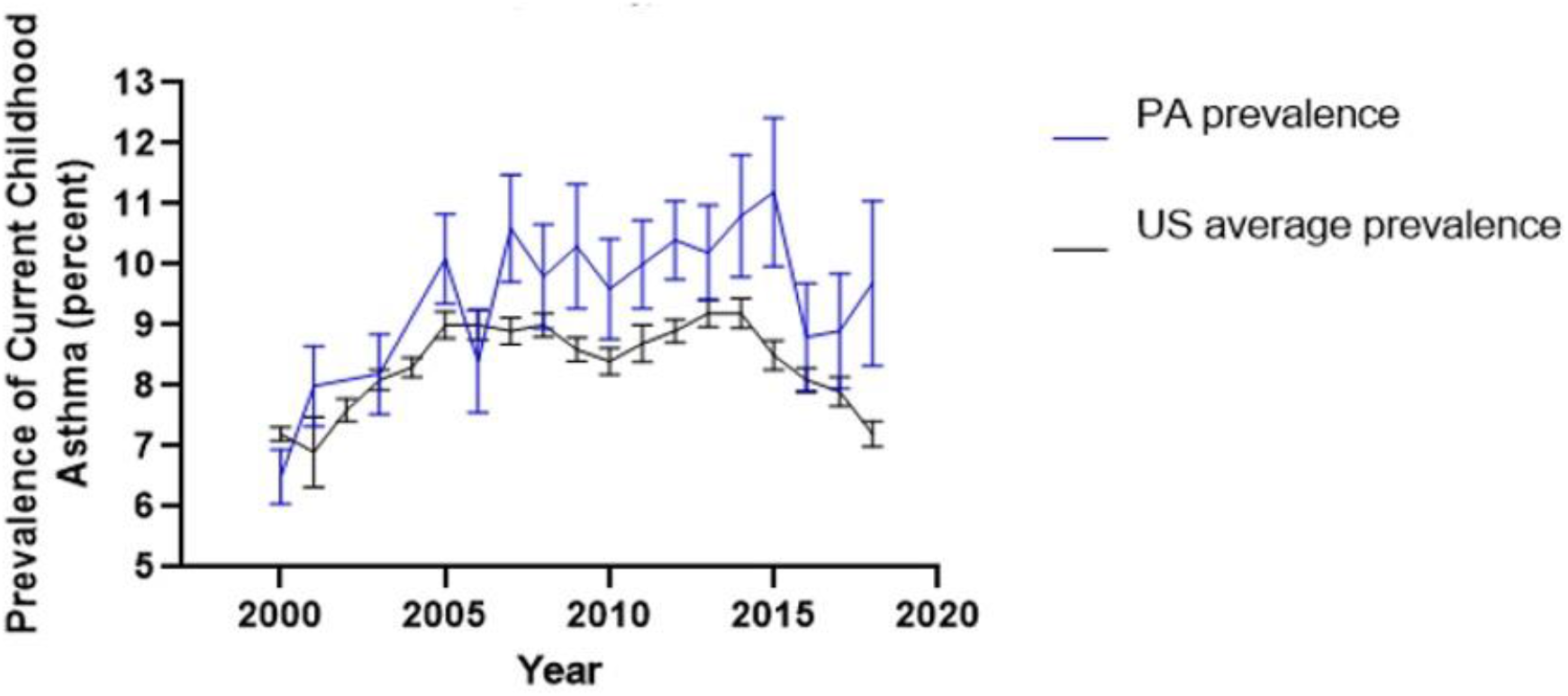
Child Asthma Prevalence in Pennsylvania (PA) compared to United States (US) average according to the CDC (17)

Figure 2 shows that approximately ¼ of people in Philadelphia have asthma, according to the 2010 report. The mean was 17.85 and the standard deviation was 4.57.

**Figure 2:**
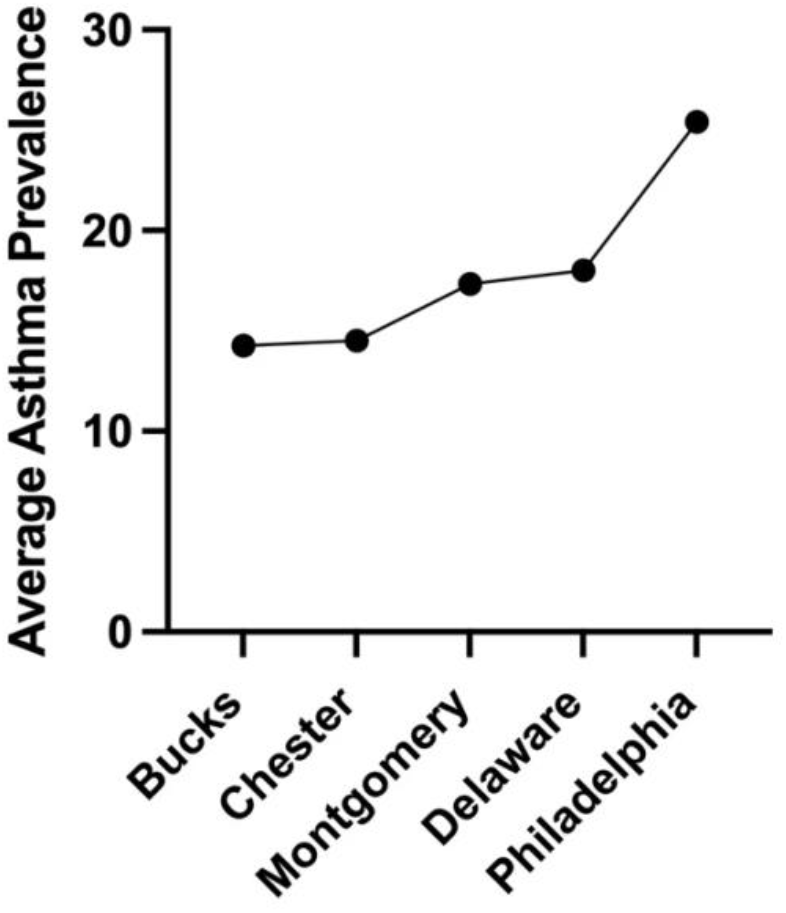
Asthma Prevalence in Surrounding Philadelphia Counties According to Community Asthma Prevention Program (21)

Figure 3 portrays the vast differences in the prevalence of asthma in the city of Philadelphia reported in 2010 from Community Health Database. Center City, Philadelphia, was found to have to highest asthma prevalence. The mean was 25.42 and the standard deviation was 6.70.

**Figure 3:**
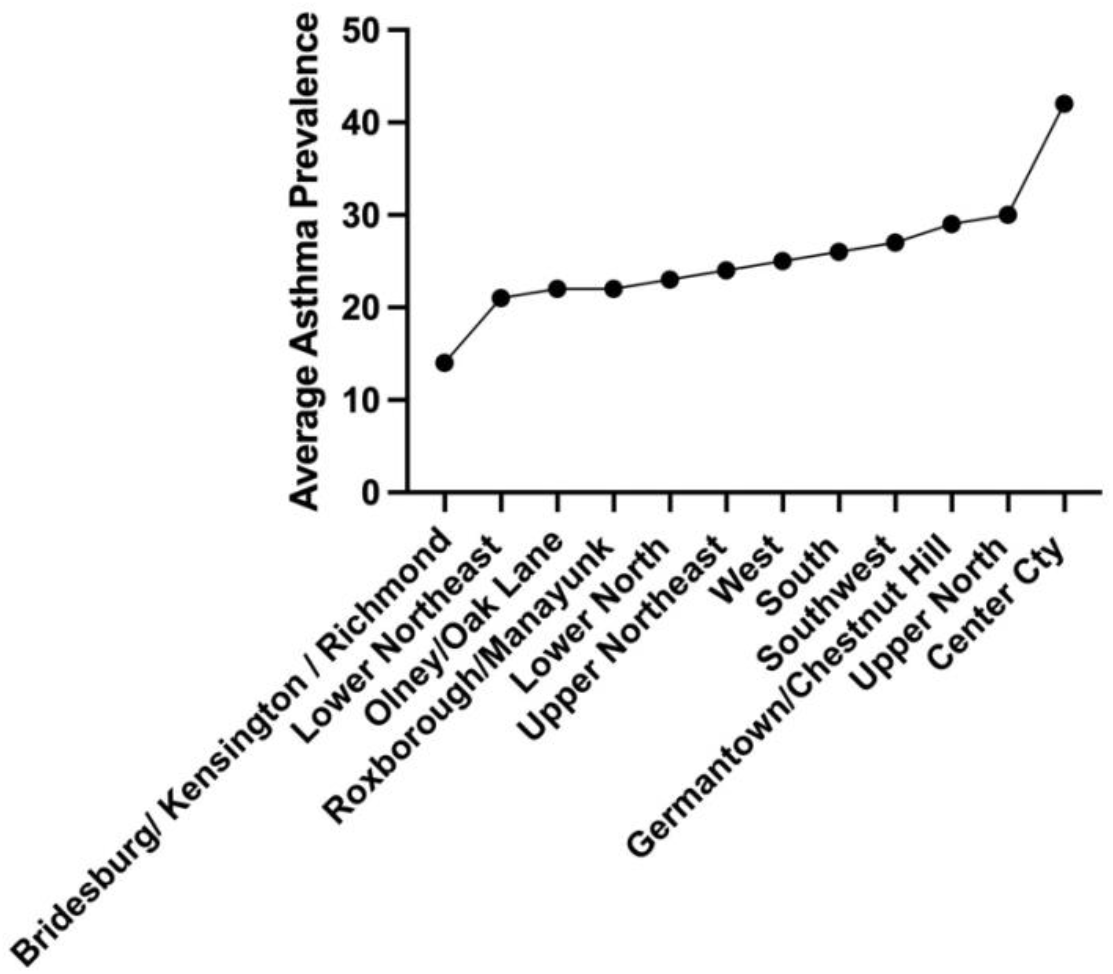
Percent of Asthma Prevalence by Philadelphia Neighborhood According to Community Asthma Prevention Program (21)

Figure 4A and 4B display the amount of confirmed asbestos damage found in tile, measured in square feet and in pipe insulation, which was measured in linear feet. The data show high amounts of reported asbestos in Carnell and Sullivan elementary schools specifically.

**Figure 4A:**
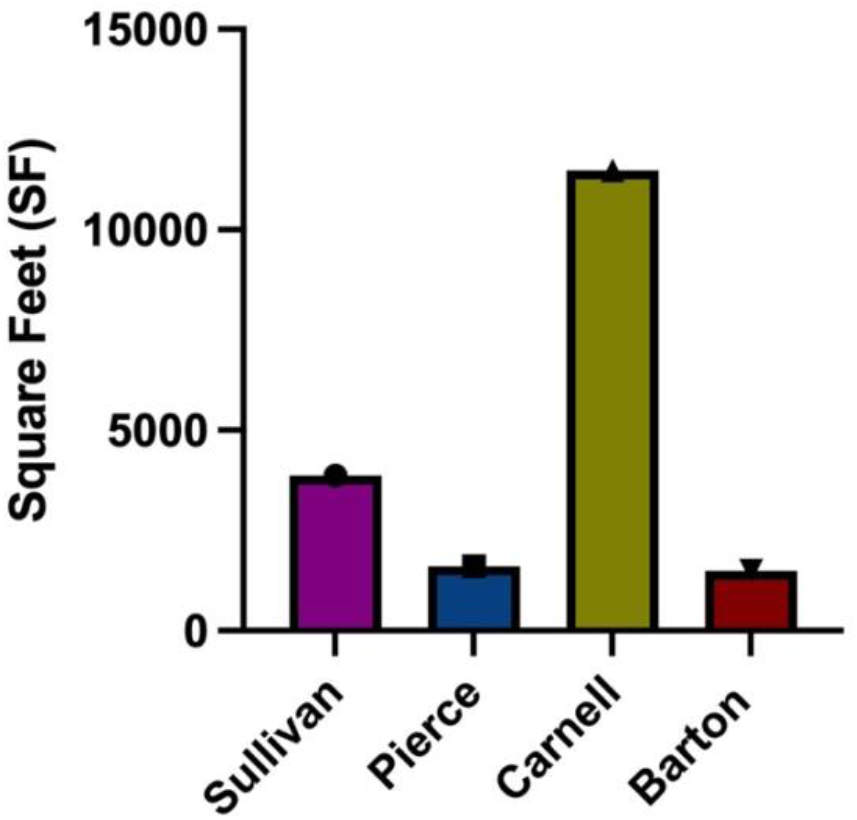
Total Asbestos Damage in Tile According to AHERA (7-10)

**Figure 4B:**
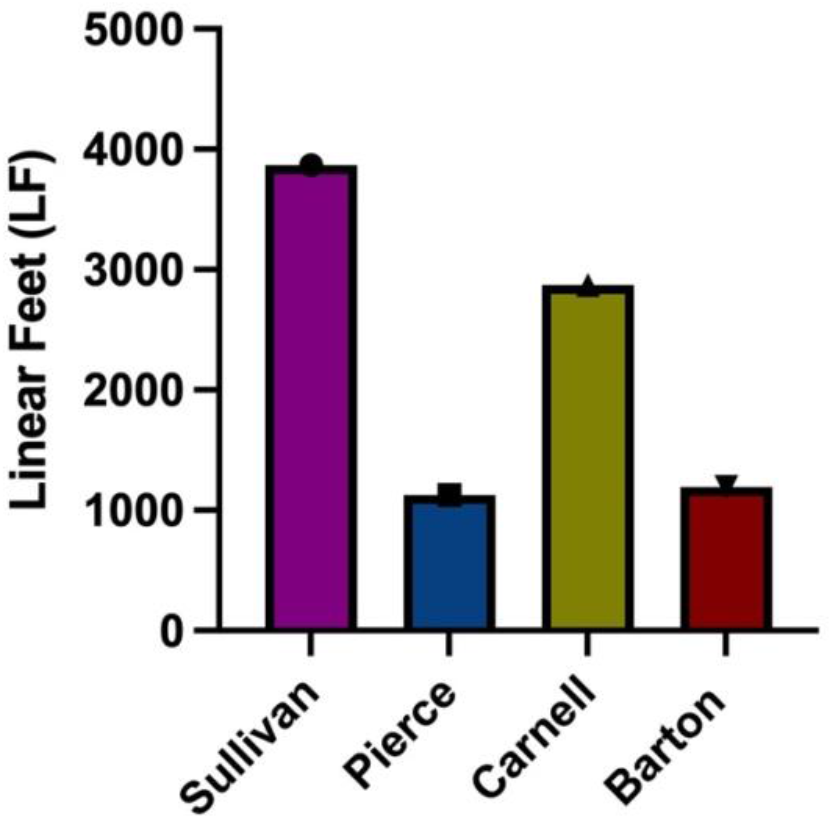
Total Asbestos Damage in Pipe Insulation According to AHERA (7-10)

Figure 5 presents the data for the amount of 2-6inch pipe insulation that had been damaged by asbestos accumulation. Pierce and Carnell elementary schools had the highest amount of pipe insulation damage.

**Figure 5:**
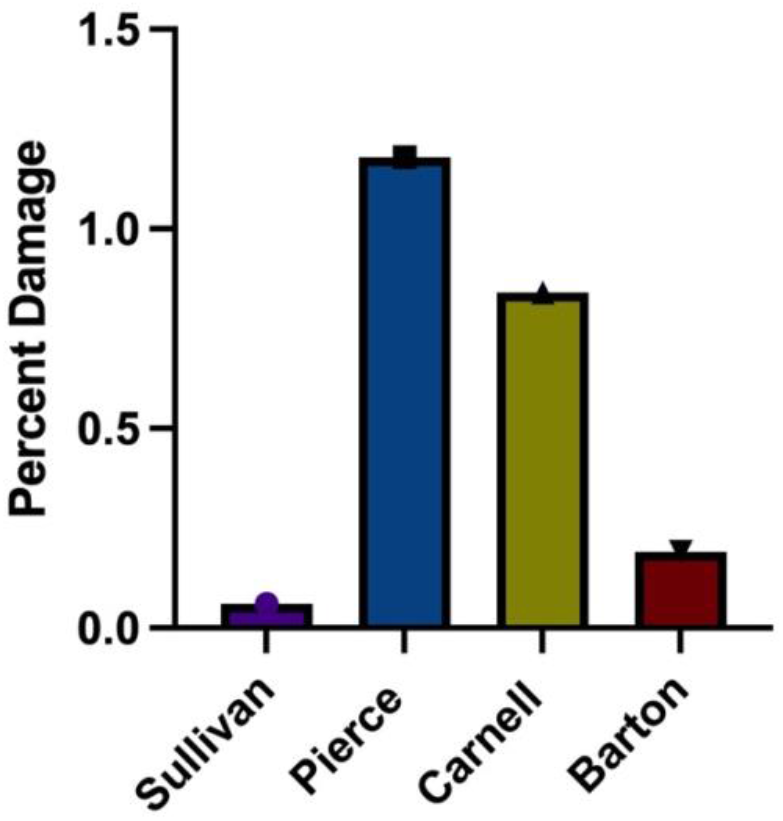
Percent Damage of Asbestos in 2-6inch Pipe Insulation According to AHERA (7-10)

Friable material is defined as an easily powdered, broken down material. This graph shows the amount of asbestos accumulation that was designated as “friable” in the AHERA reports according to the general contractor who investigated the schools.

Figure 7 illustrates the amount of asbestos and associated hazardous conditions in each of our four schools, as stated in the AHERA reports. The higher the number and letter, the worse the condition. 1/D represents undamaged asbestos, which was found in all of the schools. 6/A represents immediate concern and great risk, which occurred in Pierce elementary school.

**Figure 6:**
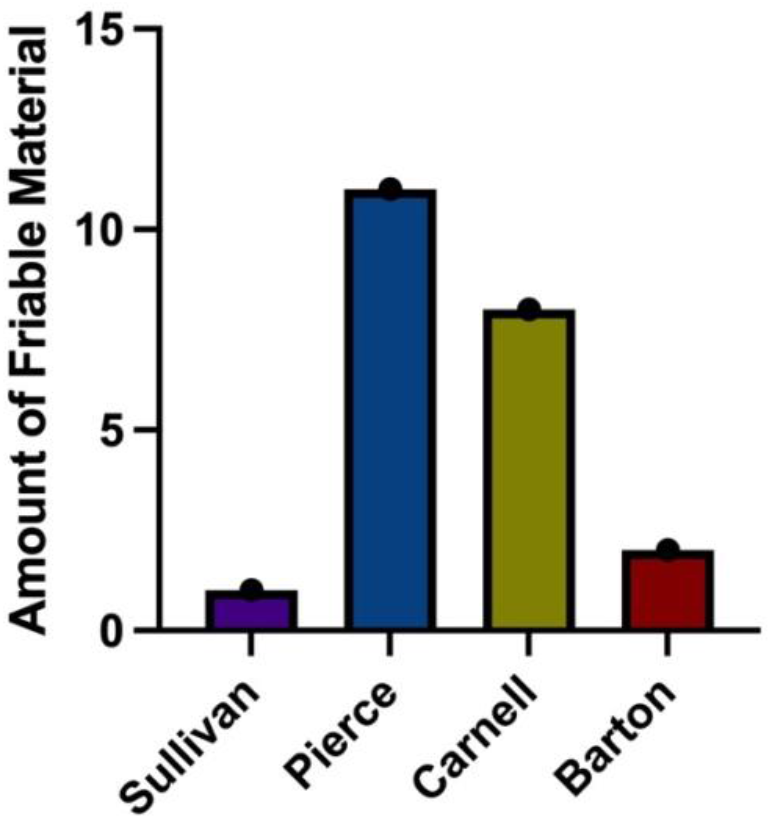
Amount of Newly Friable Material Found According to AHERA (7-10)

**Figure 7:**
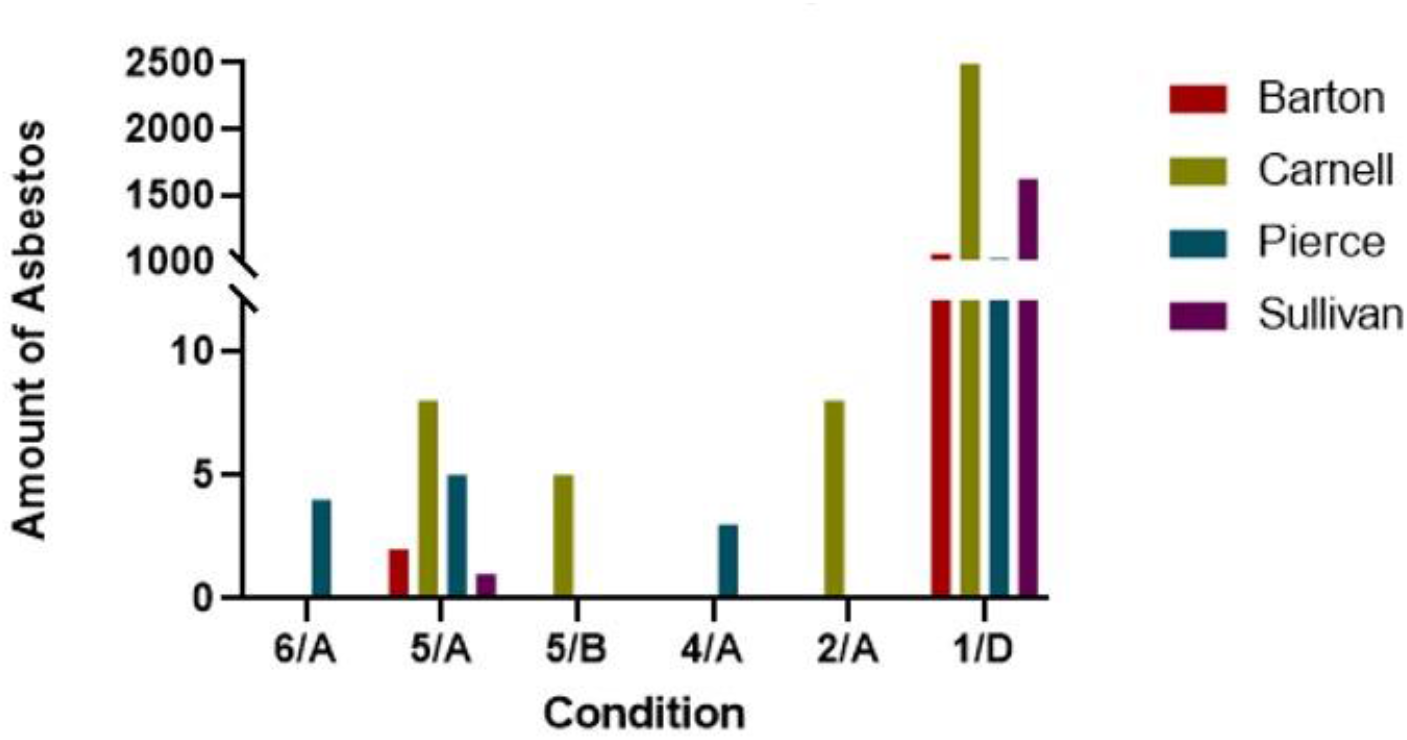
Asbestos Condition and Associated Management Plan According to AHERA (7-10)

Figure 8 portrays the data for the asthma rates for each school’s county (Pierce, Upper North; Sullivan, Lower Northeast; Carnell, Upper Northeast; Barton, Upper North) versus the newly friable material found in each school. The p value is 0.6 and the r^2^ is 0.1151.

**Figure 8:**
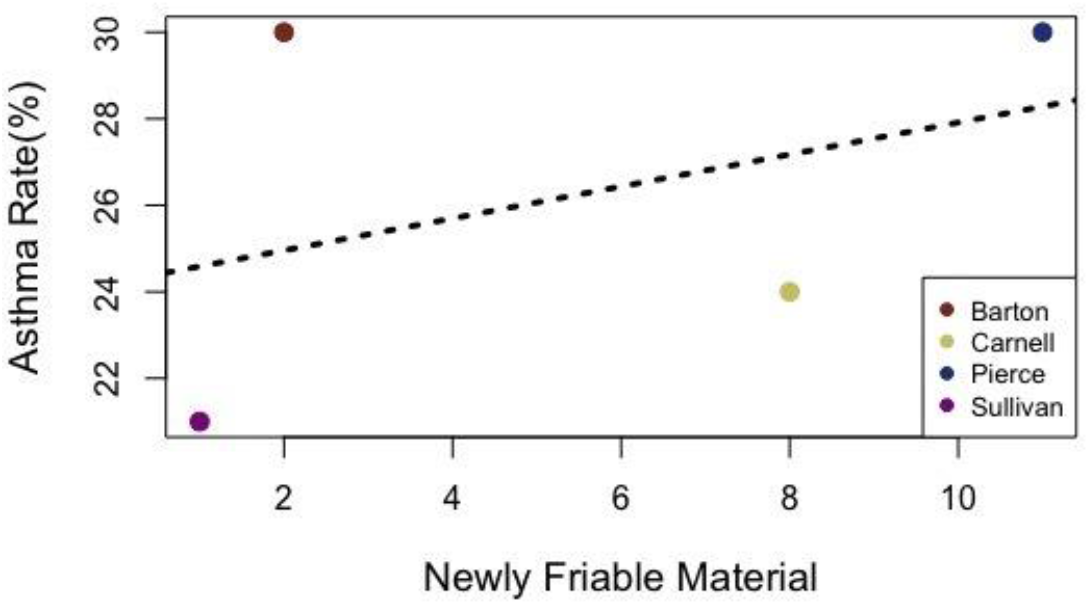
Asthma Rate in School Counties versus Newly Friable Material Found

## Discussion

The CDC reported in its Behavioral Risk Factor Surveillance System publications, that Pennsylvania has consistently had above-average rates of childhood asthma compared nationally (14). Figure 1 shows the prevalence of childhood asthma in the United States (US) and Pennsylvania (PA), as reported by the Philadelphia Department of Public Health. It can be seen that since 2000, PA has had a higher number of children with asthma compared to the rest of the nation. As of 2016, the rate is steadily inclining again in PA, while in the US it was decreasing (Figure 1). Philadelphia has a higher presence of volatile organic compounds in the city’s air. The schools analyzed that were closer to the inner city had an increase in these since 2017, according to the Philadelphia Department of Public Health (15). These particles in the air can be detrimental to those that have asthma. The risk factors that are causing the high prevalence need to be further explored and analyzed. Compared to Philadelphia County, surrounding areas have lower asthma rates, suggesting the importance of examining the factors that are influencing the rise in Philadelphia specifically (Figure 2). This showed that 25% of Philadelphia residents are diagnosed with asthma. Comparing different Philadelphia neighborhoods, it has been found that Center City residents have the highest asthma rates (Figure 3).

According to the Philadelphia Department of Public Health, Philadelphia was reported to have the highest child mortality rate compared to other United States cities, with 71.6 deaths per 100,00 children (20). Contributing conditions cited were low birth weight, prematurity, and asthma, among many others. They reported that in 2018, there were 15,450 asthma-related visits, with hospital visits being the highest among children under the age of six. This fact is also influenced by insurance status and racial/ethnic background (20). These statistics show the fundamental importance of our study.

This asbestos crisis and its effects are lasting in some communities today. Our selected Philadelphia elementary schools were chosen on the basis of having been closed as part of the 2019 asbestos crisis, to show the importance of this situation (10). Figures 4A and 4B exhibit the total amount of confirmed asbestos damage in the elementary schools, depending on the building material. There was a plethora of tile and pipe that was damaged with asbestos (Figure 4A and 4B). Carnell elementary has the most asbestos present in tile (Figure 4A), while Sullivan elementary had the most asbestos damage on pipe insulation (Figure 4B). The asbestos materials were all found in areas where students would have access to be exposed, such as classrooms and hallways. Figure 5 displays the confirmed percent damage from asbestos on 2-6 inch pipe insulation for each of the schools. Pierce Elementary school had the worst asbestos damage on this building material and this led to its eventual closure in 2019 for asbestos remediation (10) (Figure 5). The damage on the 2-6inch pipe insulation ranged from 0.06 and 1.18% (Figure 5). An important aspect of asbestos infection is the particulates breaking off and mixing into the air. The term friable is used to characterize the way the asbestos can be easily broken and crushed into powder when disturbed. This means that children who come into contact with it could easily break it down. Figure 6 illustrates the amount of asbestos damaged material designated as “friable” by the general contractor who investigated the schools and wrote the reports. Pierce elementary had the highest value of 11 friable materials (7). Interestingly, the AHERA reports designated a section to outline the conditions of the asbestos, and tagged a management plan to these conditions. Figure 7 illustrates the range of conditions associated with asbestos damage. The higher the number and letter, the worse the condition, according to the general contractor’s reports. The 1/D value can be thought of as the “watch and wait” action plan. Although it is of lowest concern, the asbestos damage is still there and needs to be managed accordingly. Peirce was the only school to contain 6/A asbestos containing material, which of the highest concern as the condition is severe (Figure 7). The children in the schools would have daily exposure to this toxic material, which could lead to respiratory aggravation in the young students. It is important that these reports were completed, because they brought awareness to the abundance of this hazardous material surrounding children. Although data analysis resulting in Figure 8 was not statistically significant, it shows the need for future development and research. With a larger sample size, the connection between newly friable material and asthma rates in each county can be better analyzed. With only four schools in our data set, this was hard to achieve. The increasing prevalence of friable material may contribute to the county’s asthma rates.

Asthma is a very serious problem, and is exceptionally bad in Philadelphia due to multiple contributing factors. It is a great concern that there is so much asbestos material in elementary schools. Younger children are at higher risk to suffer from effects from being exposed, especially those that have asthma. Philadelphia shows a higher risk for asthma than most areas of the United States and there needs to be an improvement in the public health department to help control the high rates of asthma in Pennsylvania’s children.

This study was conducted during the SARS-COVID-19 pandemic and correspondence with databases and other researchers presented challenges with slow communication. This caused gaps in our AHERA reports analysis and clarification was hard due to low correspondence with primary data collectors. Also, only having whole children population data and not age group data with the asthma rates is a limitation for this study. But the significance of this data is still important to be examined, as there are multiple factors in play that have increased these rates asthma recently.

In future studies, it would be beneficial to compile data on asthma prevalence for specific age groups, to allow for better statistical comparison. More research needs to be done on the contributing factors to increasing volatile organic compounds in Philadelphia.

## Conclusion

Asthma is relatively prevalent in Philadelphia, and may affect children through historic issues with asbestos in the schools they attend. We primarily looked at secondary data; specifically inspection data reports documenting asbestos levels in elementary schools around Philadelphia. We hope further work could include partnerships with Children’s Hospital of Philadelphia and Philadelphia Department of Health to survey data and elicit better understanding of the severity of asthma in children in the city through asbestos and air quality data. By aggregating asbestos data, the extent of the asbestos problem can be determined and show its effect on the children of Philadelphia and see how this relates to the quality of air present. This data allows future studies to examine other factors contributing to asthma in children and direct the attention for the much needed community health research.

## Supporting information

Coi Disclosure Form

## Data Availability

Data was obtained from publicly available Asbestos Hazard Emergency Response Act (AHERA) reports and publicly available data from Philadelphia Department of Public Health.

https://www.cdc.gov/asthma/brfss/default.htm

https://www.healthyschoolbuildings.com/school-data/ahera-reports/

## Acknowledgements

Shaylyn Paolello, who was a contributor at the beginning of this research project. Professor Elizabeth Kuchinski, Professors Catherine Freeland, and Dr. Brian Piper who helped advise through the research analysis process. Dr. James Church, who helped with statistical analysis. Community Asthma Prevention Program (CAPP) with Children’s Hospital of Philadelphia who gave data to aid in this report.

